# Vaccination and herd immunity thresholds in heterogeneous populations

**DOI:** 10.1101/2021.02.26.21252553

**Authors:** Elamin H. Elbasha, Abba B. Gumel

## Abstract

It has been suggested, without rigorous mathematical analysis, that the classical vaccine-induced herd immunity threshold (HIT) assuming a homogeneous population can be substantially higher than the minimum HIT obtained when considering population heterogeneities. We investigated this claim by developing, and rigorously analyzing, a vaccination model that incorporates various forms of heterogeneity and compared it with a model of a homogeneous population. By employing a two-group vaccination model in heterogeneous populations, we theoretically established conditions under which heterogeneity leads to different HIT values, depending on the relative values of the contact rates for each group, the type of mixing between groups, relative vaccine efficacy, and the relative population size of each group. For example, under biased random mixing and when vaccinating a given group results in disproportionate prevention of higher transmission per capita, it is optimal to vaccinate that group before vaccinating other groups. We also found situations, under biased assortative mixing assumption, where it is optimal to vaccinate more than one group. We show that regardless of the form of mixing between groups, the HIT values assuming a heterogeneous population are always lower than the HIT values obtained from a corresponding model with a homogeneous population. Using realistic numerical examples and parametrization (e.g., assuming assortative mixing together with vaccine efficacy of 95% and basic reproduction number of 2.5), we demonstrate that the HIT value considering heterogeneity (e.g., biased assortative mixing) is significantly lower (40%) compared with a HIT value of (63%) assuming a homogeneous population.

## 1. Introduction

Since the identification of the severe acute respiratory syndrome coronavirus 2 (SARS-CoV-2) and subsequent devastating impact of the 2019 novel coronavirus pandemic (COVID-19), a pertinent question being asked by the global public health and scientific research community has been what is the minimum fraction of the susceptible population that needs to be immunized for the pandemic to end? It has been widely reported that the herd immunity threshold (HIT) vaccination coverage, denoted by *v*^*^, is given by the formula 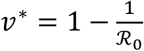, where ℛ is *the basic reproduction number* of the model, defined as the average number of secondary infections caused by an infective person over his or her infectious period when introduced into a completely susceptible population [1]. For example, ℛ_0_ = 2.5 indicates the need to immunize more than 60% of the population to achieve HIT, so that the disease becomes eliminated from the population. However, this derivation for HIT is based on making several assumptions regarding the properties of the vaccine and characteristics of the population. Specifically, it assumes that the vaccine efficacy to protect against the acquisition of infection is perfect and lasts throughout the lifetime of the vaccinated individual (i.e., the vaccine does not wane).

McLean and Blower [2], and other researchers [3,4], derived modified HIT formulae under various assumptions for vaccine properties. Their derivation is based on considering the following scenario for a cohort (childhood) vaccine. Suppose a hypothetical vaccine is efficacious in a fraction, *ε*, of recipients and confers full protection that wanes at a rate *ω per* unit time in a population with average life expectation of 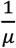, so that the average duration of vaccine-induced protection is 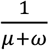. In this case, the critical vaccination coverage (i.e., the proportion of newborns that need to be vaccinated to achieve herd immunity, consequently leading to disease elimination), denoted by *v*^**^, is given by [3]

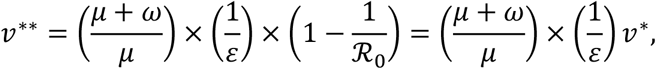

where *v*^*^ is as defined above. Thus, the herd immunity threshold (*v*^*^) needs to be adjusted upward to reflect the imperfect efficacy of the vaccine (0 < *ε* < 1) and the fraction of the average lifetime a vaccinated individual remains protected (1/(*μ* + *ω*) ÷ 1/*μ*). For example, with ℛ_0_= 2.5 and a vaccine with protective efficacy of only 70% (i.e., *ε* = 0.7) that lasts 90% of a lifetime (i.e., 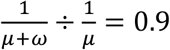) is used in the population, at least 95.2% of the population of the unvaccinated susceptible newborns needs to be vaccinated to achieve disease elimination (i.e., achieve HIT), since *v*^**^ = 0.952 in this case.

The above derivations assume a homogeneous, well-mixed population. However, the transmission of many infections (such as SARS-CoV-2) occurs in a diverse heterogeneous population. Hence, a more realistic approach to carry out the above computations will be to account for the relevant heterogeneities [5]. In other words, the computations need to be carried out for the case where the total population is divided into multiple groups with similar characteristics, such as age, contact patterns, infectious period, or social, cultural, demographic, or geographic factors. For example, several mathematical models for disease transmission employ different mixing patterns, such as those between different age groups [6,7]. These contact patterns are typically parametrized using empirical or synthetic social contact matrices estimated from population-based surveys [8].

## 2. Formulation of Disease Transmission Model in Heterogeneous Populations

The multigroup vaccination model for the transmission dynamics of a disease (such as SARS-CoV-2) in a heterogeneous population typically follows a standard susceptible-vaccinated-exposed-infected-recovered (SVEIR) Kermack-McKendrick-type compartmental modeling formulation [7, 9]. In this formulation, the total population at time t (denoted by *N*(*t*)) is sub-divided into *m* distinct homogeneous groups. Each group is further sub-divided into five disjoint (mutually exclusive) classes or compartments of unvaccinated susceptible (*S*(t)), vaccinated susceptible (*V*(*t*)), exposed (*E*(*t*)), infectious (*I*(*t*)), and recovered/removed (*R*(*t*)), so that *N*_*i*_ = *S*_*i*_ + *V*_*i*_ + *E*_*i*_ + *I*_*i*_ + *R*_*i*_, with *i* = 1,2,…, *m*. Mathematically speaking, ‘exposed individuals’ are those who are newly infected with the pathogen but are not yet able to transmit the pathogen to other individuals (i.e., they are not infectious yet) [7].

The resulting SVEIR model, for the transmission dynamics of a disease in *m* heterogeneous groups or populations, is given by the following deterministic system of nonlinear differential equations (where a dot represents differentiation with respect to time *t*):

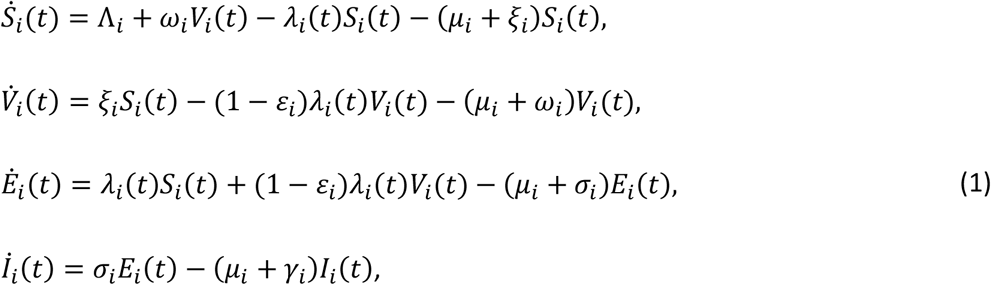

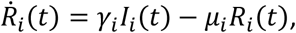

where the *force of infection λ*_*i*_(*t*) is given by

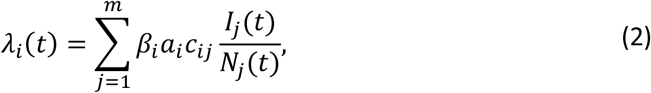

with *β*_*i*_ as the transmission probability *per* contact for group *i, a*_*i*_ is the average number of contacts that an individual of group *i* has during a certain period of time (called group-specific activity level), and *c*_*ij*_ is the proportions of contacts that members of group *i* have with other individuals of group *j*. Mixing should meet the following closure relation [10]:

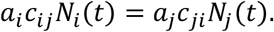

That is, the total number of contacts that individuals of group *i* have with other individuals of group *j* during a certain period of time should be equal to the total number of contacts that individuals of group *j* have with other individuals of group *i*. In the model (1), heterogeneity between groups is captured through differences in demographic rates (i.e., birth and death rates), transmission probability per contact, contact rates, progression and recovery rates, and vaccine efficacy and waning rates.

Adding all the equations of the model (1) gives the following equation for the rate of change of the total population:

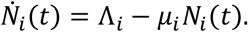

In the model (1), Λ_*i*_ is the *per capita* recruitment (birth) into the population, *ω*_*i*_ is the vaccine waning rate, *λ*_*i*_(*t*) is the force of infection, *μ*_*i*_ is the natural death rate (i.e., 1/*μ*_*i*_ is the average lifespan of a person in group *i*) and *ξ*_*i*_ is the vaccination rate. Furthermore, 0 < *ε*_*i*_ < 1 is the protective efficacy of the vaccine, *σ*_*i*_is the rate at which exposed individuals develop clinical symptoms of the disease (i.e., 1/*σ*_*i*_ is the latency period of the disease), and γ_*i*_ is the recovery rate.

Some of the main assumptions made in the formulation of the model (1) are:

a. Within-group homogeneous mixing (i.e., although the model considers *m* heterogeneously mixed groups, contact patterns within each group is homogeneous).
b. Exponentially distributed waiting time in each epidemiological compartment.
c. The vaccine is not perfect (i.e., 0 < *ε*_*i*_ < 1), and the protection offered by the vaccine wanes over time (i.e., *ω* > 0). In addition, the vaccine has no therapeutic benefits.
d. No disease-induced mortality (so that the total population in each group remains constant).
e. Recovery induces permanent immunity against acquisition of future infection.

### 2.1 Disease-free equilibrium of general model in heterogeneous populations

The model (1), for a single group, has been subjected to rigorous mathematical analysis in the literature. Specifically, results for its well-posedness, invariance of its solutions, and existence and asymptotic stability of equilibria (disease-free and endemic) have been established [7, 11]. The multigroup model

(1) has a unique disease-free equilibrium given by

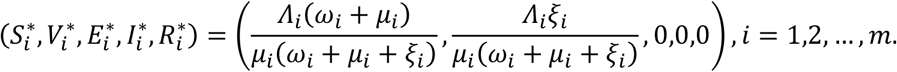

It is convenient to assume that the population in each group *i* has reached a stationary (equilibrium) state such that

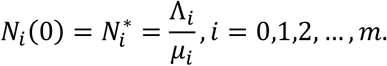

It is also convenient to work with the fraction of the population in each group. The proportion of individuals in group *i* that are vaccinated (at the disease-free equilibrium) is given by:

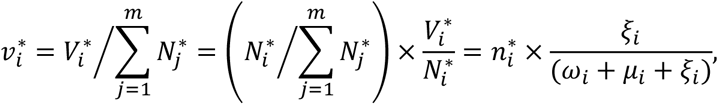

where,

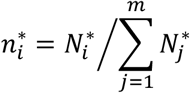

is the fraction of total individuals in group *i* relative to total population.

## 3. Analysis of Model in Heterogeneous Populations with Two Risk Groups

Here, we focus on deriving analytic expressions for HITs for a special case of the heterogeneous population model (1) that considers only two risk groups (i.e., the model (1) with *m* = 2). For a two-group model, we first derive the vaccination reproduction number, ℛ_*v*_, and then find the minimum proportion of individuals in the community that need to be vaccinated in order to reduce ℛ_*v*_ to a *v*alue less than 1, so that herd immunity is achieved.

### 3.1 Vaccination and basic reproduction numbers for two-group model

The next-generation operator method [13] can be used to compute the vaccination reproduction number (and, subsequently, the basic reproduction number) of the special case of the model (1) with m = 2. It follows, using the notation in [12], that the non-negative matrix of new infection terms (*F*) and the M-Matrix (*V*) of linear transition terms in the infected compartments are given, respectively, by (where 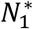 and 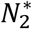 are the total population of group 1 and group 2, respectively; similarly, 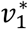 and 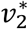 represent the HIT for groups 1 and 2, respectively):

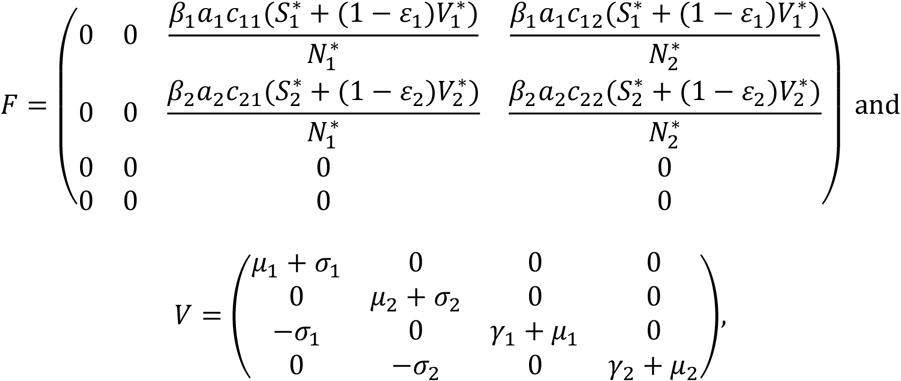

from which it follows that the vaccination reproduction number is given by (where *ρ* is the spectral radius)

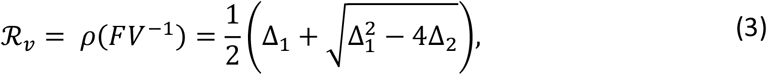

where,

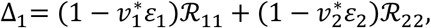

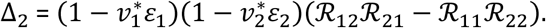

In deriving equation (3), we utilized the following definition of the constituent basic reproduction numbers associated with disease transmission between individuals in group *i* with individuals in group *j*:

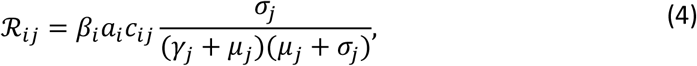

where the index *i* represents group *i* and the index *j* represents group *j*. To obtain the basic reproduction number (ℛ_0_) associated with the two-group model, we set the vaccination coverage rates in the expression for ℛ_*v*_, given by (3), to zero (i.e., we set 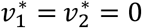 in (3)). This gives,

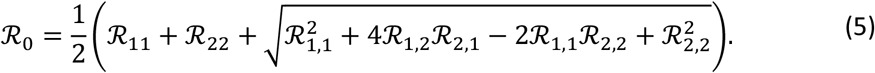

### 3.2 Herd immunity thresholds for a two-group model with heterogeneous populations

For herd immunity threshold determination or computation of a two-group model, the objective is to find the values of the respective HITs, 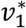 and 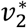, such that the total vaccine coverage (i.e., the proportion of individuals in the community that is vaccinated), given by

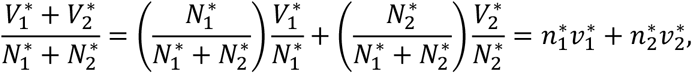

is at its minimum and vaccinated reproduction number, ℛ_*v*_, given by (3), is less than or equal to one. Formally, the optimization problem can be written as choosing 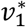 and 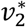 to

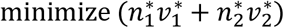

subject to,

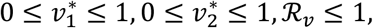

where ℛ_*v*_ is given by equation (3). The solution of this nonlinear optimization problem will be characterized using a geometrical approach. Specifically, we compare the shape of the curve depicting values of vaccination coverage where the vaccinated reproduction number is equal to one (ℛ_*v*_ = 1) with the contour lines (or level sets) 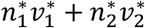 (as illustrated in Figure 1). Each contour line represents the locus of vaccination coverage combinations 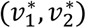 that yield the same level of total vaccination coverage at the population level. The blue contour lines correspond to lower and lower levels of total vaccination coverage when moving in the southwestern direction toward the origin.

**Figure 1.**
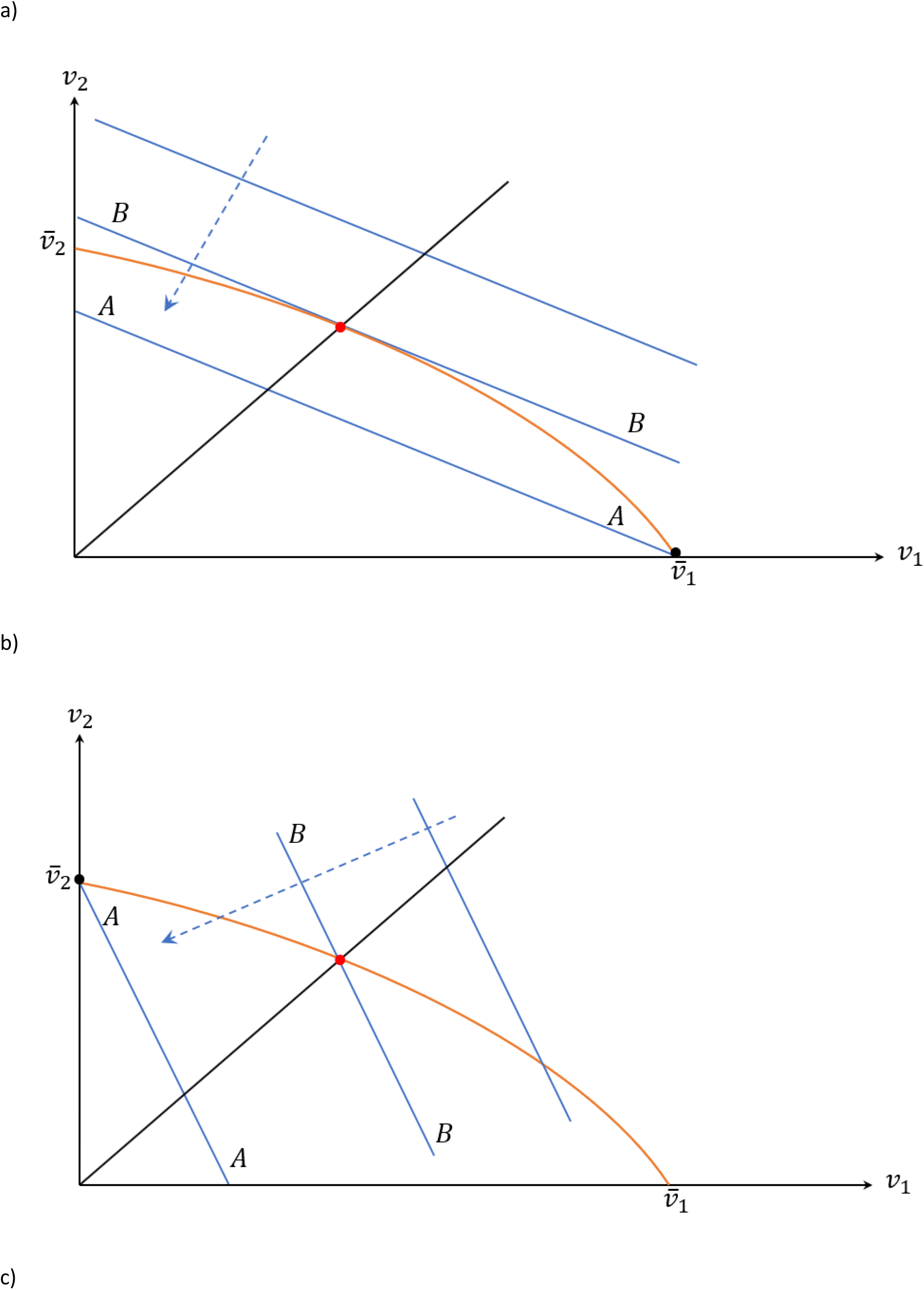
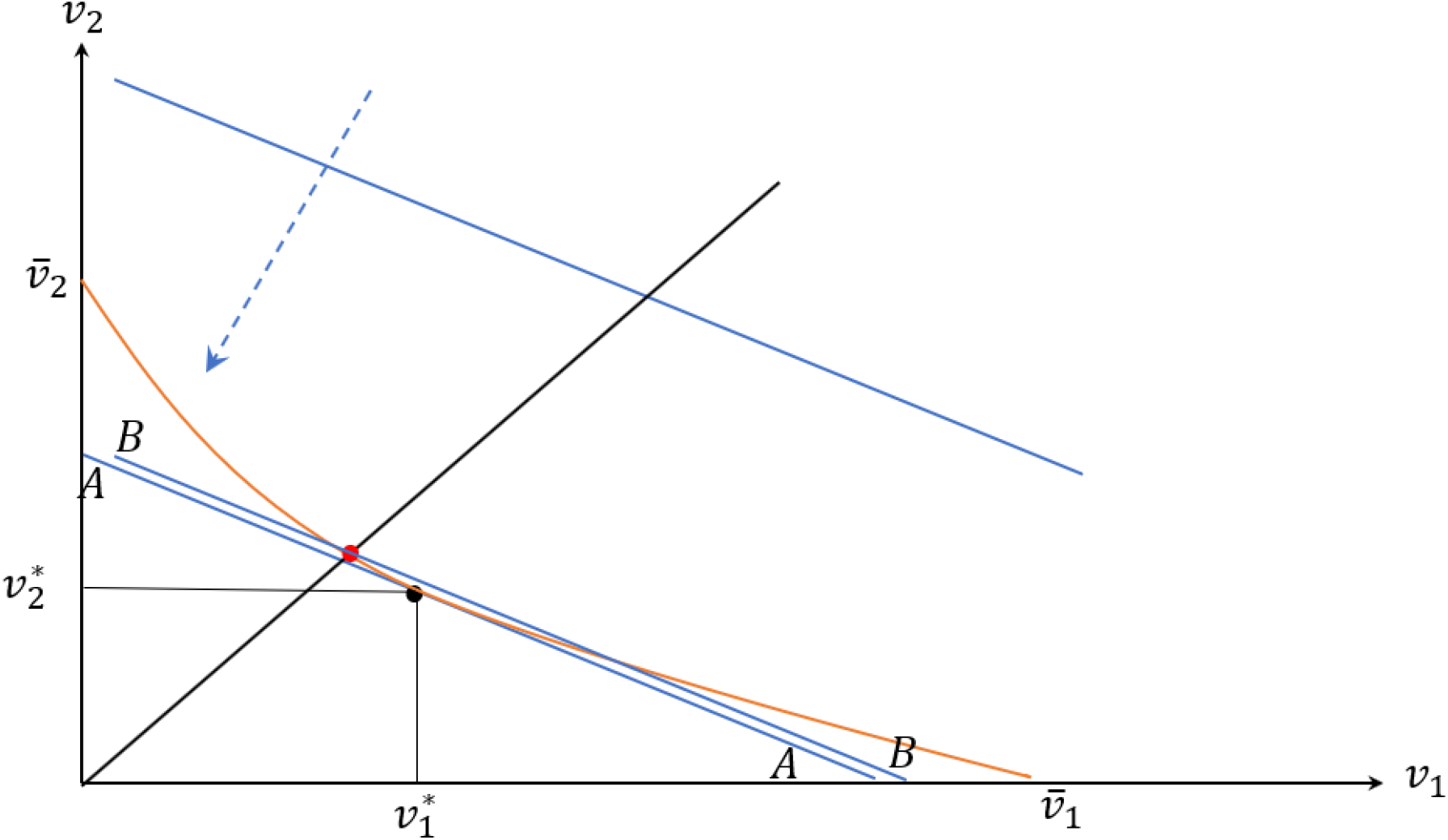
Vaccination threshold values for group 1 and group 2. The orange curve shows values of vaccination coverage where the vaccinated reproductive number is equal to one: ℛ_*v*_ = 1. The slope of this curve when intersects with the *y-*axis is given by 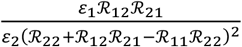. The blue level curves show different values of total vaccination coverage going down in the direction of the origin: 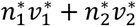. The slope of these level curves is – 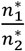. Under biased random mixing (a) when the blue line is flatter than the orange curve when it intersects the *y-*axis, the closest blue line to the origin that intersect the orange curve happens when 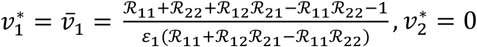: (b) when the blue line is steeper than the orange curve when it intersects the *x-* axis, the closest blue line to the origin that intersect the orange curve happens when 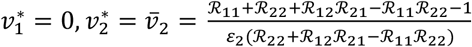. Under biased assortative mixing (c) the optimum occurs when the blue line is tangent to the orange curve. In the homogeneous population model 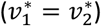, the optimum occurs at the intersection of the black 45-degree line with the orange curve.

The solution of the equation of the orange curve ℛ_*v*_ = 1 yields a value of 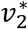 as a function of 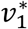:

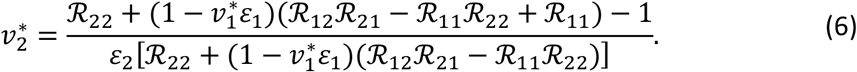

It follows from equation (6) that this function intersects the *x-*axis 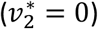 and *y-*axis 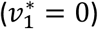 at

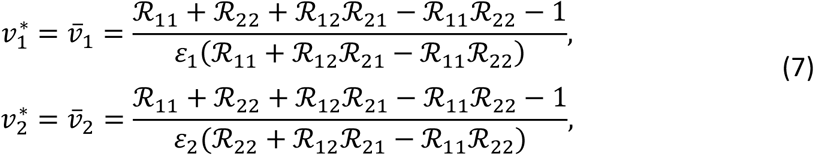

respectively. The slope of the orange curve (Figure 1) is given by

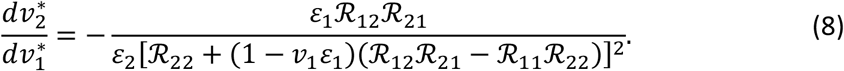

The slope of the orange curve, at the point of intersection with the *y-*axis (i.e., 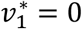), is given by

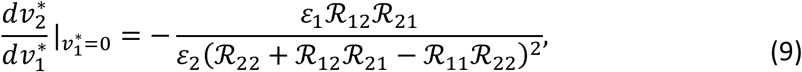

and the slope of the orange curve when it intersects with the *x-*axis (i.e., 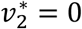) is given by

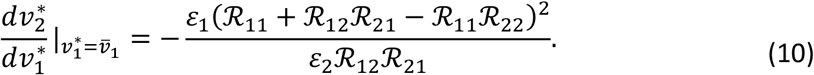

The equations of the blue level curves are: 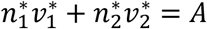, where *A* is an arbitrary constant (Figure 1).

The slope of these level curves is

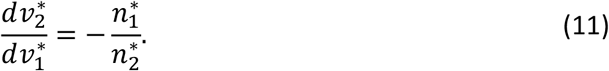

Depending on the situation, the geometrical approach involves comparing the slopes of the curves given by equations (8)–(11). We consider the following two scenarios, based on which group(s) the vaccine is prioritized to.

### Scenario 1: Vaccinating only one group

We start with the case where ℛ_*v*_ can be brought down to one by vaccinating either group 1 alone or group 2 alone. That is, the values of 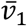 and 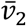 given in equation (7) are less than 1.

We note that the type of mixing between groups (i.e., *c*_*ij*_) determines the sign of the expression ℛ_12_ℛ_21_ − ℛ_11_ℛ_22_. By using equation (4), we can establish that

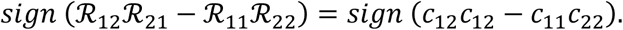

We distinguish two types of mixing.

#### A) Biased random mixing: ℛ_12_ℛ _21_ ≥ ℛ_11_ℛ _22_

When transmission occurs more because of mixing between groups rather than because of mixing within the groups, we call this type of mixing *biased* toward *random mixing*. The *separable proportionate mixing* is a special type of biased random mixing [7, 10].

i) Vaccinating group 1 disproportionately contributes more to prevention of *per-capita* transmission. Here, the following inequality holds:

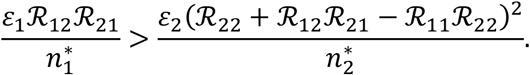

Figure 1a depicts the situation where the absolute value of the slope of the orange curve when it intersects the *y*-axis (equation (9)) is greater than the absolute value of the slope of the blue line (equation (11)) (i.e., blue line is flatter). That is (hence the inequality above),

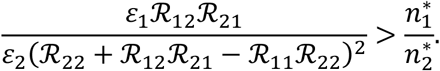

In this case, the closest blue line to the origin that intersect the orange curve happens when

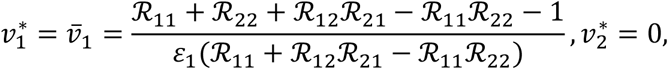

and the herd immunity threshold is given by

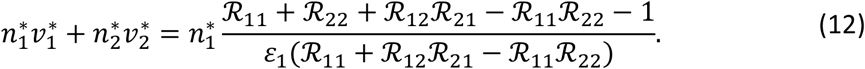

(ii) Vaccinating group 1 disproportionately contributes less to prevention of per-capita transmission:

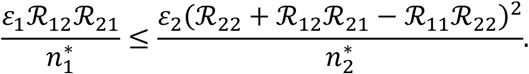

In this case, the absolute value of the slope of the orange curve when it intersects the y-axis (equation (9)) is less than or equal the absolute value of the slope of the blue line (equation (11)) (i.e., blue line is steeper) (illustrated in Figure 1b),

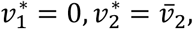

and the herd immunity threshold is

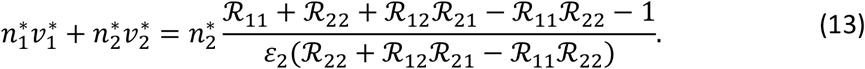

In summary, the above results show that, under the biased random mixing assumption, achieving herd immunity entails exclusively optimizing vaccination coverage among the group that results in relatively more prevention of *per-capita* disease transmission.

#### B) Biased assortative mixing: ℛ_12_ℛ _21_ < ℛ_11_ℛ _22_

When transmission occurs more due to mixing within groups rather than due to mixing between groups, we refer to this type of mixing as *biased* toward *assortative mixing*. In this case, the optimization problem has both boundary (corner) and interior solutions. We start with the two boundary solutions followed by the interior solution.

i) Vaccinating group 1 disproportionately contributes more to prevention of *per-capita* transmission:

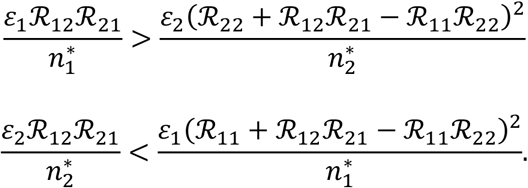

In this case, the absolute values of the slope of the orange curve when it intersects the *y-*axis (equation (9)) and *x-*axis (equation (10)) are greater than the absolute value of the slope of the blue line (equation (11)) (i.e., blue line is flatter). In this case,

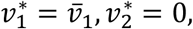

and the herd immunity threshold is given by equation (12). In other words, we achieve optimal results by allocating all necessary vaccine resources to group 1.

ii) Vaccinating group 1 disproportionately contributes less to prevention of *per-capita* transmission. Here, we have:

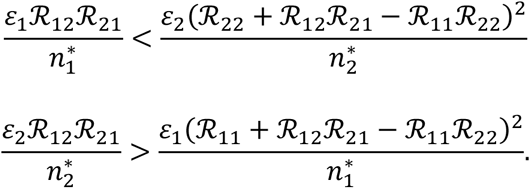

In this case the absolute values of the slope of the orange curve when it intersects the *y-*axis (equation (9)) and *x-*axis (equation (10)) are less than the absolute value of the slope of the blue line (equation (11)) (i.e., blue line is steeper). In this case,

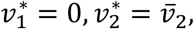

and the herd immunity threshold is given by equation (13). Here, optimal results are achieved by allocating all vaccine resources to group 2.

### Scenario 2: Vaccinating both groups

This scenario includes one interior solution and 2 boundary solutions. For this scenario, we do not need to assume that ℛ_*v*_ can be brought down to one by vaccinating either group 1 alone or group 2 alone.

i) Interior solution. For this solution to occur, the following inequalities must hold:

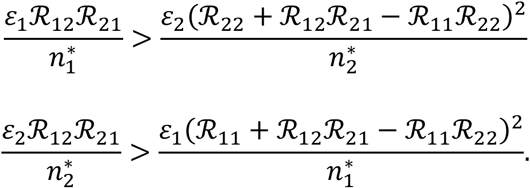

When the absolute value of the slope of the blue line (equation (11)) is between the absolute values of the slope of the orange curve when it intersects the *y-*axis (equation (9)) and *x-*axis (equation (10)), it is optimal to vaccinate both groups (illustrated in Figure 1c). The interior solution is obtained when the slope of the orange curve (equation (8)) is equal to the slope of the blue curve (equation (11)). Thus, by equating the right-hand side of equations (8) and (11) and solving for 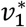 and using the resulting solution in equations (6), we have:

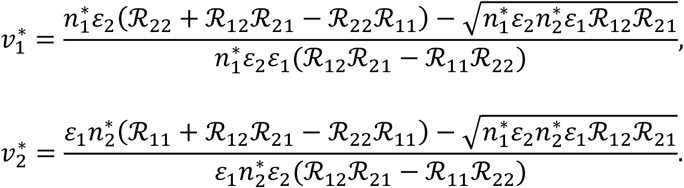

It follows from the assumption of biased assortative mixing and the inequalities above a positive **interior** solution exists if

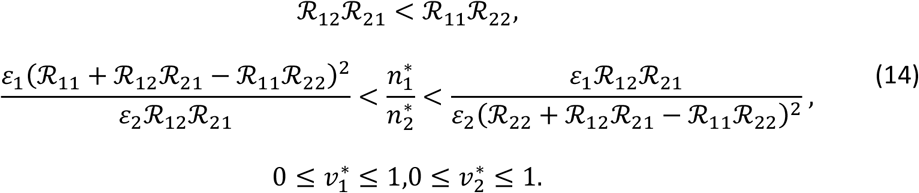

The herd immunity threshold is

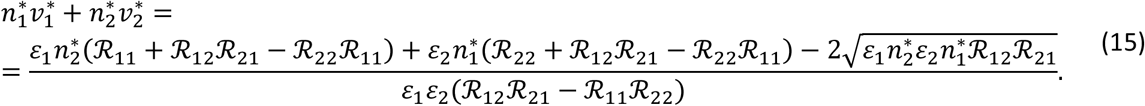

ii) Boundary solutions.

The case where ℛ_*v*_ cannot be brought down to one by vaccinating either group 1 alone or group 2 alone (i.e., the values of either 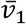 or 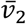 are greater than 1) always leads to boundary solutions. For example, when 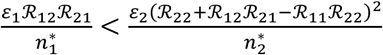 and 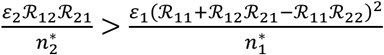 and 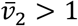, we first maximize vaccination coverage among group 2 and then find the value of coverage among group 1 that brings ℛ_*v*_ to one. In this case,

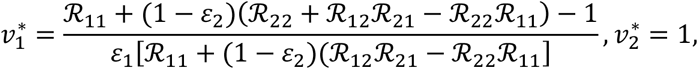

and the herd immunity threshold is

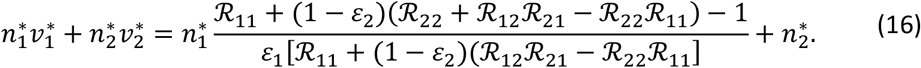

Similarly, when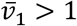, we may have,

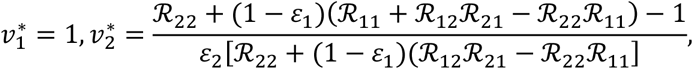

and the herd immunity threshold is

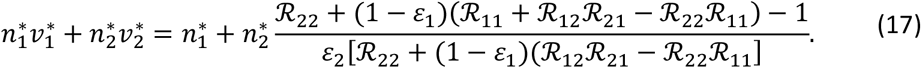

Thus, the optimal solution and HIT are summarized as follows:

1. an interior solution if inequalities given in (14) are satisfied. The corresponding HIT value is given by equation (15);
2. four boundary solutions exist if any of inequalities given in (14) does not hold. The associated HIT value is given by equations (12), (13), (16), and (17);
3. no solution otherwise.

These analyses show that, for a two-group vaccination model in heterogeneous populations, the optimum vaccination program depends on the relative values of the constituent reproduction numbers, ℛ_12_, ℛ_21,_ℛ_11_, ℛ_22_, relative vaccine efficacy, and the relative population sizes of the two groups. The values of the constituent reproduction numbers are determined by the type of mixing allowed or assumed between the two groups. When the mixing between the groups is biased towards random mixing, achieving herd immunity entails restricting vaccination coverage to the group that results in relatively more prevention of *per-capita* transmission. If herd immunity cannot be achieved by vaccinating one of the two groups alone, vaccination coverage among the group that results in relatively more prevention of *per-capita* transmission should be maximized before vaccinating the other group.

These scenarios occur under biased assortative mixing too, but scenarios involving vaccinating both groups are more common, including interior optimum where coverage of any of the two groups is less than 100%.

## 4. Analysis of Model with a Homogeneous Population

The model (1) can be reduced to that with homogeneous population by assuming that each individual in any of the two groups is identical with every other individual in the community. That is, we achieve homogeneity by setting *c*_*ij*_= *c, a*_*i*_ = *a, ξ*_*i*_ = *ξ, ε*_*i*_ = *ε, μ*_*i*_ = *μ*, ⋀_*i*_ = ⋀, *σ*_*i*_ = *σ*, γ_*i*_ = γ for all *i* and *j* into the model (1). Specifically, the vaccination model with a homogeneous population is obtained from system (1) by dropping the group subscript *i* and re-defining the force of infection as

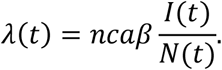

Using the next-generation operator method [13], the *vaccination reproduction number* associated with the resulting homogeneous model, denoted by ℛ_*v*_, is given by:

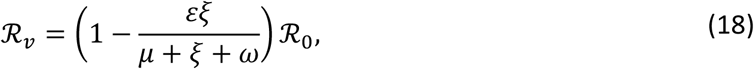

where the basic reproduction number, ℛ_0_, is given by

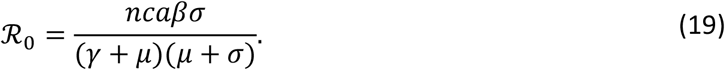

Since the vaccination coverage at the disease-free equilibrium (*v*^*^) for the multigroup model with homogeneous population is

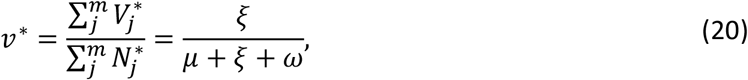

it follows, by using (18) in (20), that

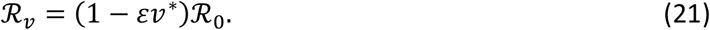

It should be noted that this relationship between ℛ_*v*_ and ℛ_0_ (equation (21)) in the homogeneous population model can obtained directly from the formula of the vaccination reproduction number of the heterogeneous population model by substituting *ε*_*i*_ = *ε* and 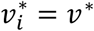, *i* = 1,2, into equation (3) and using the definition of ℛ_0_ given by equation (5).

Setting ℛ_*v*_ to one and solving for *v*^*^ gives the following threshold value needed to achieve herd immunity for the model with homogeneous population [3, 7]:

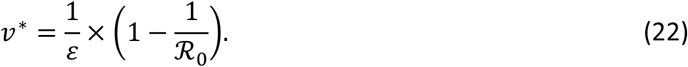

Thus, in constructing the homogeneous population model corresponding to the heterogeneous model (1) for comparison, we followed previous studies and matched the two modeling types (i.e., homogeneous vs. heterogeneous population) according to the expressions or values of their respective basic reproduction number, ℛ_0_ [14,15].

### 4.1 Comparison of HIT values using homogeneous and heterogeneous population models under proportionate mixing

One of the simplest types of mixing in disease epidemiology is the separable proportionate mixing, in which the contacts of a person of group *i* are distributed over those of other groups in proportion to the activity levels and sizes of the other groups [7]. Thus, with proportionate mixing, proportions of contacts that members of group *i* have with group *j, c*_*ij*_, is given by

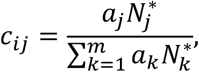

where *a*_*i*_ and 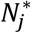 are as defined before.

By substituting this definition of *c*_*ij*_ into the formulae for the constituent reproduction numbers in equation (4), it can be shown that the assumption of proportionate mixing implies that ℛ_12_ℛ_21_ = ℛ_11_ℛ_22_, which in turns implies, Δ_2_= 0. Thus, for this scenario of proportionate mixing, the vaccination reproduction number reduces to,

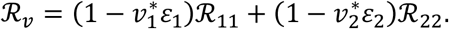

We now seek to answer the question: what are the values of 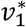 and 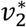 such that total vaccination coverage, 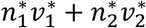, is at its minimum and *R*_*v*_ ≤ 1? The answer depends on the relationship between the ratio of constituent reproduction numbers 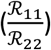 adjusted by efficacy and the ratio ofpopulation 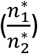 in the two groups. Solution of this simple linear programming problem is a special case of the biased random mixing where ℛ_12_ℛ_21_ = ℛ_11_ℛ_22_. As before, there are three scenarios:

**Scenario (i)** Vaccinating group 1 disproportionately contributes more to prevention of *per-capita* transmission: 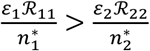.

In this case,

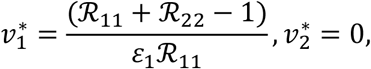

and the herd immunity threshold is

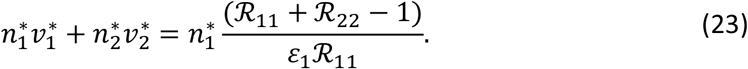

**Scenario (ii)** Vaccinating group 1 disproportionately contributes less to prevention of *per-capita* transmission: 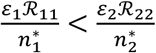.

In this case,

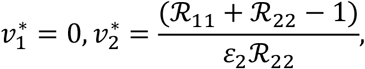

and the herd immunity threshold is

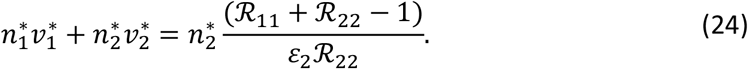

**Scenario (iii)** Vaccinating both groups contribute equally to prevention of transmission: 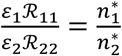 In this case, values of 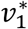 and 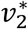 such that

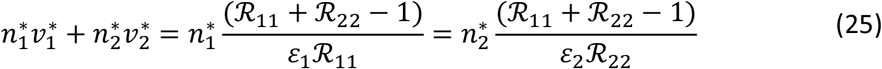

will yield the minimum fraction that need to be vaccinated to achieve herd immunity.

To facilitate the comparison of the herd immunity thresholds between homogeneous and heterogeneous population models, we need to make sure efficacy in the two models is the same. One approach is to assume that efficacy does not vary across the two groups such that *ε*_1_ = *ε*_2_ = *ε*.

If 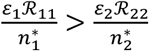, the threshold vaccine coverage under heterogeneous population model is (given by the right-hand side of equation (23)):

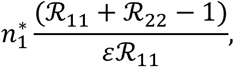

and that for the homogeneous population model is given by

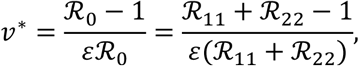

since ℛ_0_ = ℛ_11_ + ℛ_22_ > 1 under proportionate mixing.

It can be shown that

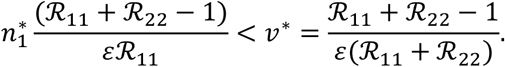

Upon simplifications, the above inequality holds if

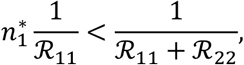

Noting (and using) our starting assumption 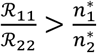, it follows that 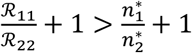 or

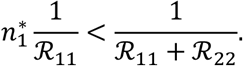

Therefore, it follows from the above inequality, that the threshold vaccine coverage in the heterogeneous population model, under scenario (i), is always less than that in the corresponding homogeneous population model.

Similarly, if 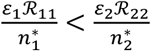, we can follow the same approach above and show that the threshold vaccine coverage under heterogeneous population model, under scenario (ii), given by equation (24) is always lower than that under the homogeneous population:

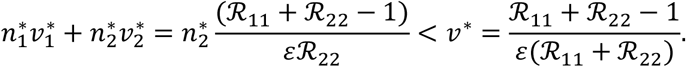

When 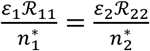, it follows, under equal efficacy, that 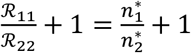 or

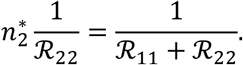

Thus, the threshold vaccine coverage under heterogeneous population model, under scenario (iii), given by equation (25) is always equal to that under the homogeneous population:

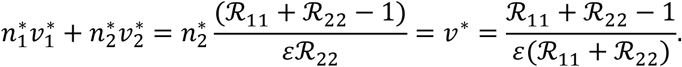

Therefore, under the form of proportionate mixing between groups, we show analytically that the HIT value in the heterogeneous population model is always less than or equal to the HIT value in a homogeneous population model.

### 4.2 Comparison of HIT values using homogeneous and heterogeneous population models under general mixing

To show that the HIT value in the heterogeneous population model is always less than or equal to the HIT value in a homogeneous population model under general mixing between groups we utilize the geometrical approach depicted in Figure 1. Recall that the blue level curve represents total vaccination coverage. Thus, line *AA* corresponds to HIT value in heterogeneous population model whereas *BB* corresponds to HIT value in the homogeneous population model. We consider scenarios with two boundary solutions and one interior solution:

i. Vaccinating group 1 only (Figure 1a). In this case, line *AA* is closer to the origin than line *BB*. Hence, the HIT value in heterogeneous population model is lower than the HIT value in the homogeneous population model.
ii. Vaccinating group 2 only (Figure 1b). In this case, line *AA* is closer to the origin than line *BB*. Hence, the HIT value in heterogeneous population model is lower than the HIT value in the homogeneous population model.
iii. Vaccinating both group 1 and group 2 (Figure 1c). In this case, line *AA* is closer to the origin than line *BB*. Hence, the HIT value in heterogeneous population model is lower than the HIT value in the homogeneous population model.

## 5. Numerical Analysis of HIT Values in Heterogeneous and Homogeneous Population Models

Figure 2 illustrates numerically the different scenarios leading to different HIT values in the heterogeneous population model and compare them with the HIT values in a corresponding homogeneous population model. The orange curve shows values of vaccination coverage where ℛ_*v*_ = 1 and the blue level curves show different values of total vaccination coverage going down in the direction of the origin 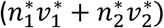. In a homogeneous population model, vaccination coverage (determined by the intersection with the orange curve) is uniform across the two groups as shown by the dotted black 45-degree line. In all scenarios we set vaccine efficacy to 95% across the two groups.

**Figure 2.**
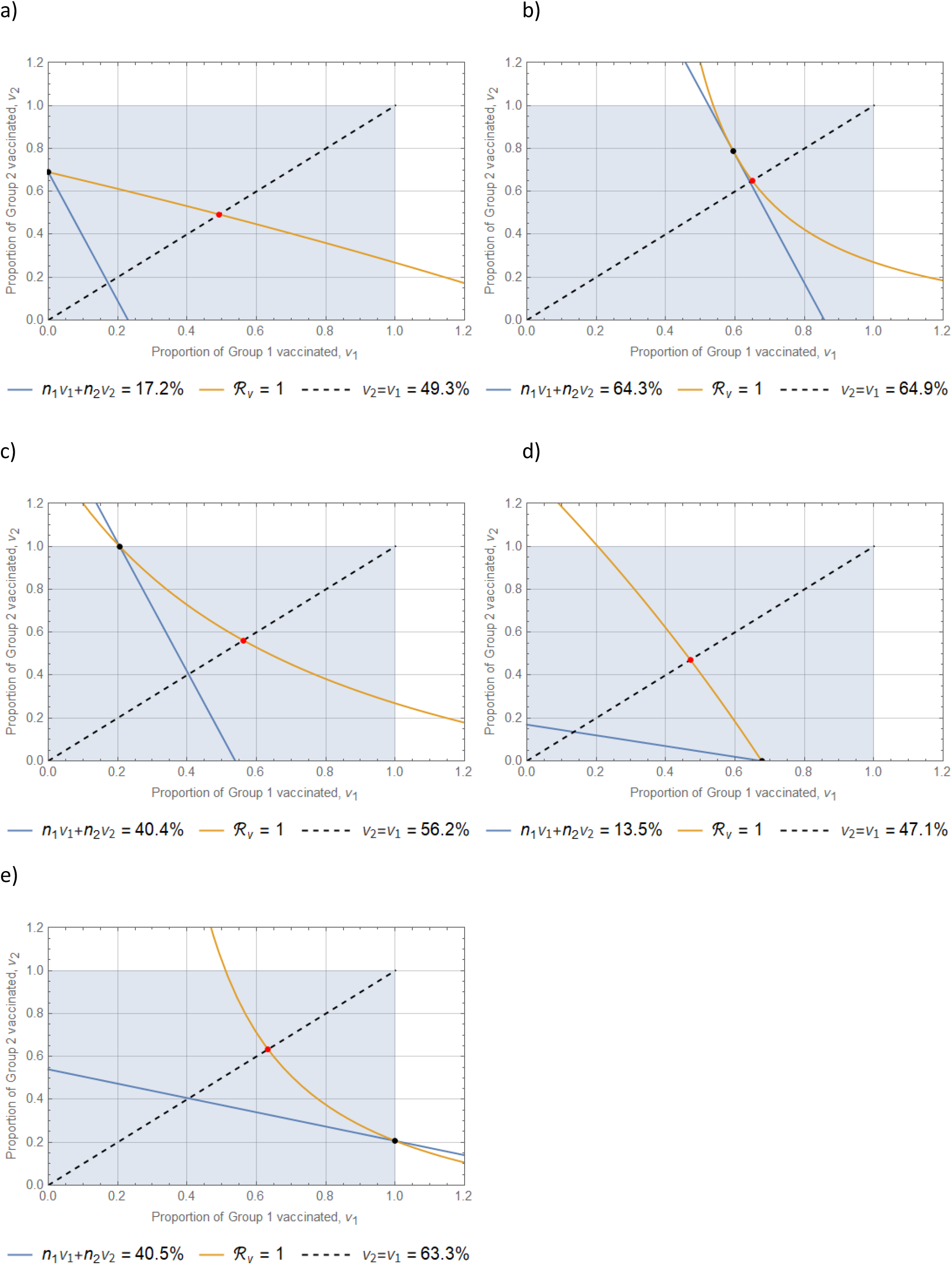
Illustration of optimal least vaccine coverage determination by group that satisfies the constraint ℛ_*v*_ = 1 (orange curve). Parameter values: *ε* = 0.95, *ε*_1_ = 0.95, *ε*_2_ = 0.95. (a–c): ℛ_12_ = 1.0, ℛ_21_ = 0.8, ℛ_22_ = 1.3, 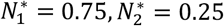. a) ℛ_11_ = 0.5, ℛ_0_ = 1.88; b) ℛ_11_ = 2, ℛ_0_ = 2.6; c) ℛ_11_ = 1.2, ℛ_0_ = 2.16; d) ℛ_12_ = 0.8, ℛ_21_ = 1.0, ℛ_22_ = 0.5, 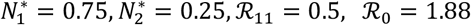; e)ℛ_12_ = 0.8, ℛ_21_ = 1.0, ℛ_22_ = 0.5, 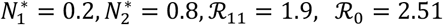.

a. The chosen parameters values represent a situation of biased random mixing between groups, and 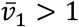 and 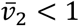. As a result, the blue line is steeper than the orange curve when the latter intersects the *y-*axis, and the closest blue line to the origin that intersect the orange curve happens when 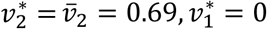 (Figure 2a). Given that group 2 represents only 25% of the population, the overall HIT value is just 17.2% compared with the HIT value in a homogeneous population model of 49.3%.
b. The chosen parameters values represent a situation of biased assortative mixing between groups, 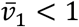 and 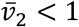, and conditions (14) is satisfied. As a result, it is optimal to vaccinate both groups 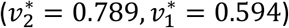 for an overall HIT value of 64.3% (Figure 2b). The HIT value in a homogeneous population model is 64.9%.
c. The chosen parameters values represent a situation of biased assortative mixing between groups, and 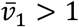 and 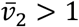. As a result, the blue line is steeper than the orange curve, and it is optimal to vaccinate all of group 2 and 20.6% of group 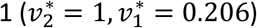 for an overall HIT value of 40.4% (Figure 2c). The HIT value in a homogeneous population model is 56.2%.
d. The chosen parameters values represent a situation of biased random mixing between groups and 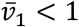 and 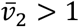. As a result, the blue line is flatter than the orange curve, and it is optimal to vaccinate group 1 only 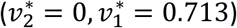 for an overall HIT value of 13.5% (Figure 2d). The HIT value in a homogeneous population model is 47.1%.
e. The chosen parameters values represent a situation of biased assortative mixing between groups and 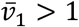 and 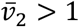. As a result, the blue line is flatter than the orange curve, and it is optimal to vaccinate all of group 1 and 20.6% of group 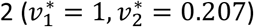 for an overall HIT value of 40.5% (Figure 2e). The HIT value in a homogeneous population model is 63.3%.

Figure 3 illustrates different optimal solutions and HIT values for various values of the basic reproduction numbers. The figure shows the situation where relative to its small size group 2 is contributing more to transmission for low values of ℛ_11_and ℛ_0_ and there is a need to vaccinate more people from group 2. As ℛ_11_(and ℛ_0_) increases, necessary vaccination coverage among group 2 increases until all of group 2 is vaccinated. As ℛ_11_(and ℛ_0_) increases further, both groups are vaccinated, but vaccination coverage among group 1 increases whereas that among group 2 decreases. Of note, the herd immunity threshold for the homogeneous population (red curve) is consistently higher, but the difference between the two shrinks as the basic reproduction number increases.

**Figure 3.**
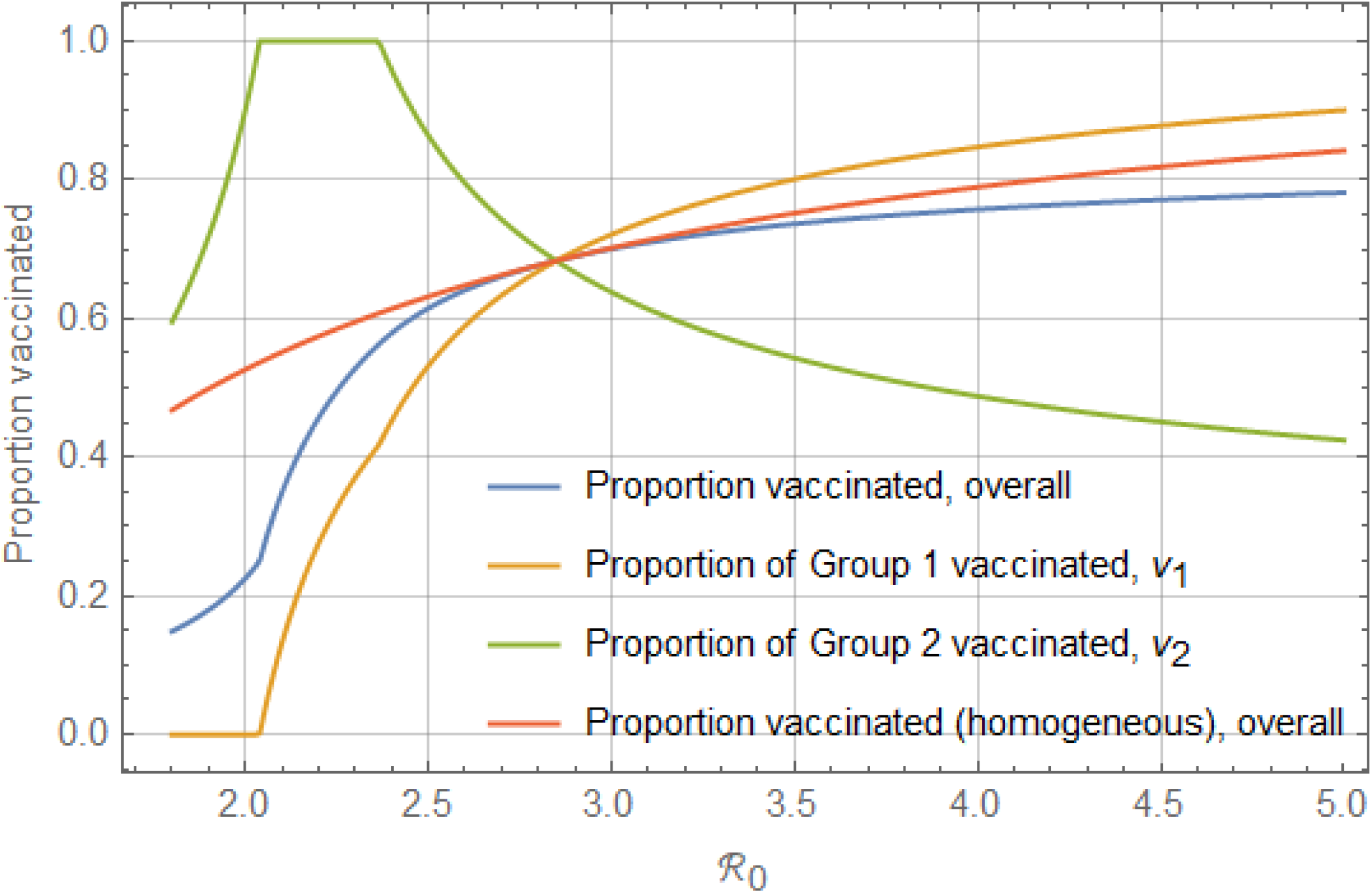
Vaccine coverage above which herd immunity is achieved by group and basic reproduction number. Parameters values: ℛ_12_ = 1.0, ℛ_21_ = 0.8, ℛ_22_ = 1.3, *ε* = 0.95, *ε*_1_ = 0.95, *ε*_2_ = 0.95, 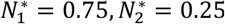. Using the definition of ℛ_0_, we chose 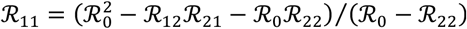.

## 6. Discussion

Our theoretical results, for a two-group model, suggest that deriving the vaccine-induced herd immunity threshold in the heterogeneous population model is much more complicated than for the model with homogeneous population. The herd immunity threshold for each vaccinated group depends on the relative values of the constituent reproduction numbers, relative vaccine efficacy, and the relative population sizes of the two groups. The values of the reproduction numbers are determined by the level and duration of infectiousness of a contact for each group, contact rates for each group, as well as the type of mixing between groups. We show that, under biased random mixing and when vaccinating a given group results in disproportionately prevention of higher transmission *per capita*, it is optimal to optimize vaccination of that group before vaccinating other groups. We also found situations, under biased assortative mixing assumption, where it is optimal to vaccinate more than one group.

We show that population heterogeneities tend to result in lower HIT values, compared with the homogeneous population case. This is true under both proportionate and other types of mixing among heterogeneous populations. Using realistic numerical examples and parametrization (e.g., assuming biased assortative mixing with vaccine efficacy of 95% and basic reproduction number of 2.5) we illustrate this finding, where the HIT value considering heterogeneity is shown to be significantly lower (40%) compared with a HIT value assuming a homogeneous population (63%).

Although our analysis utilized a two-group model, our findings can be extended to models of multiple groups. Although more complicated than a two-group model, rigorous analyses of more realistic models with many heterogeneous groups can be conducted using the methods included this paper.

## Data Availability

The work is theoretical. No new data has been generated.

## Acknowledgements

ABG acknowledges the support, in part, of the Simons Foundation (Award #585022) and the National Science Foundation (Award #1917512).

